# Localized prebiotic nitrate supplementation remodels oral biofilm metabolism and reduces gingival inflammation: a randomized placebo-controlled trial

**DOI:** 10.64898/2026.04.22.26351516

**Authors:** Brianne Yi, Hee Yeon Kim, William Sotka, Robert Estey, Shawn J Green, Harlan J. Shiau

**Affiliations:** Department of Advanced Oral Sciences and Therapeutics, University of Maryland School of Dentistry, Baltimore, Maryland, USA; Per Os Bioscience, Maryland, USA; MyFitStrip LLC, Rockville, Maryland, USA; Lundquist Institute, UCLA Harbor, Torrance, California, USA

## Abstract

**Background:** Gingival inflammation is associated with dysbiotic oral biofilms characterized by reduced nitrate-reducing capacity and diminished nitric oxide (NO) bioavailability. While dietary nitrate has been shown to influence oral microbial activity, the effects of sustained, localized nitrate delivery on oral biofilm ecology and gingival inflammation remain incompletely defined.

**Methods and findings:** In this randomized, double-blind, placebo-controlled trial, 30 adults with gingival bleeding were assigned to receive localized prebiotic nitrate (∼0.989 mmol per dose) or placebo for 21 days. The primary outcome was mean bleeding on probing (mBOP). Secondary outcomes included modified Gingival Index (mGI), Quigley–Hein plaque index (QHPI), salivary nitrite (as a proxy for NO bioavailability), oral pH, and microbiome composition assessed by 16S rRNA gene sequencing.

Nitrate supplementation significantly reduced mBOP (25.7% to 15.3%; p = 0.0002) compared to placebo. Salivary nitrite levels and oral pH increased, indicating enhanced nitrate metabolism. Microbiome analysis demonstrated enrichment of nitrate-reducing taxa, including *Rothia mucilaginosa* and *Neisseria* spp., and a relative reduction in inflammation-associated genera such as *Prevotella* and *Porphyromonas*. No significant differences were observed in plaque index, consistent with functional modulation of the biofilm rather than reduction in plaque accumulation.

**Conclusions:** Localized prebiotic nitrate supplementation was associated with reduced gingival inflammation and shifts in oral microbiome composition consistent with enhanced nitrate-reducing capacity critical in nitric oxide formation. These findings support a role for biofilm-directed nutritional modulation as a non-antimicrobial approach for managing gingival inflammation and improving nitric oxide bioavailability.

## Introduction

Gingival inflammation, commonly assessed by bleeding on probing (BOP), is a hallmark of active periodontal disease and reflects ongoing host–microbe interactions at the tooth–gingiva interface.^1,2^ Periodontal diseases, encompassing gingivitis and periodontitis, affect an estimated 20–50% of the global population and remain a leading cause of oral morbidity worldwide.^3^ Rather than being driven by a single pathogen, gingival inflammation arises from ecological disruption of the oral biofilm, characterized by a shift from health-associated commensals toward anaerobic, proteolytic, and pro-inflammatory taxa that provoke sustained host immune activation and tissue damage.^4^

Consequently, strategies that restore biofilm function—rather than broadly suppress microbial load—have emerged as a promising frontier in periodontal disease prevention and management.^5^ One such strategy involves modulation of the oral nitrate–nitrite–nitric oxide (NO) pathway.^6-8^ Dietary nitrate, abundant in leafy greens, root vegetables, such as beets, as well as inorganic nitrate from potassium salt, acts as a prebiotic substrate for facultative anaerobic oral bacteria, most notably species within the genera *Rothia* and *Neisseria*.^6–17^ These organisms reduce nitrate to nitrite, which can be subsequently converted to NO within the oral cavity. NO is a pleiotropic signaling molecule with well-established antimicrobial, vasodilatory, and anti-inflammatory properties, capable of shaping the local biochemical environment and influencing both microbial ecology and host responses.^14-27^ Importantly, nitrate metabolism selectively favors nitrate-reducing commensals while inhibiting NO-sensitive, inflammation-associated taxa, including *Fusobacterium nucleatum, Prevotella melaninogenica, Porphyromonas gingivalis*, and certain *Veillonella* species.^11,14,16-26^

Clinical studies using nitrate-rich foods or beetroot juice have demonstrated increases in salivary NO, shifts toward a more eubiotic oral microbiome, and reductions in gingival inflammation. ^16-26^ However, systemic dietary approaches are frequently constrained by variability in intake, palatability, gastrointestinal tolerance, and limited nitrate substantivity in the oral cavity. Localized delivery strategies that prolong oral nitrate availability may therefore offer greater precision and efficacy. Functional chewing gum or slow-release nitrate lozenge represents an attractive platform for such local delivery, as it enables sustained, low-dose nitrate exposure or ‘high-time’ directly within oral biofilms while enhancing contact with resident nitrate-reducing communities.^21^

Here, we conducted a randomized, double-blind, placebo-controlled trial to evaluate whether a formulated prebiotic nitrate chewing gum can reduce gingival inflammation and remodel oral biofilm composition. We hypothesized that daily localized nitrate delivery would attenuate gingival inflammation, increase salivary NO and oral pH, and promote a microbial shift toward nitrate-reducing commensals. To test this, we assessed clinical outcomes alongside biochemical markers and characterized microbiome changes using 16S rRNA gene sequencing.

## Results

### Study pool description

A total of 30 subjects were enrolled in this clinical study, with individuals randomly allocated to the control (n=15) and test (n=15) group (**Figure 1**). No subjects discontinued participation due to adverse events. Of the individuals completing the study there were 20 female and 10 males, ranging in age from 22-64 (mean 28.9 years old). One salivary plaque sample at the baseline timepoint was lost in transport, thus reducing available microbial analysis samples from 15 to 14 for the test group. There were no significant differences in overall age, race or ethnicity composition between the subjects of the enrolled control and test cohorts (**Table 1**).

**Figure 1.**
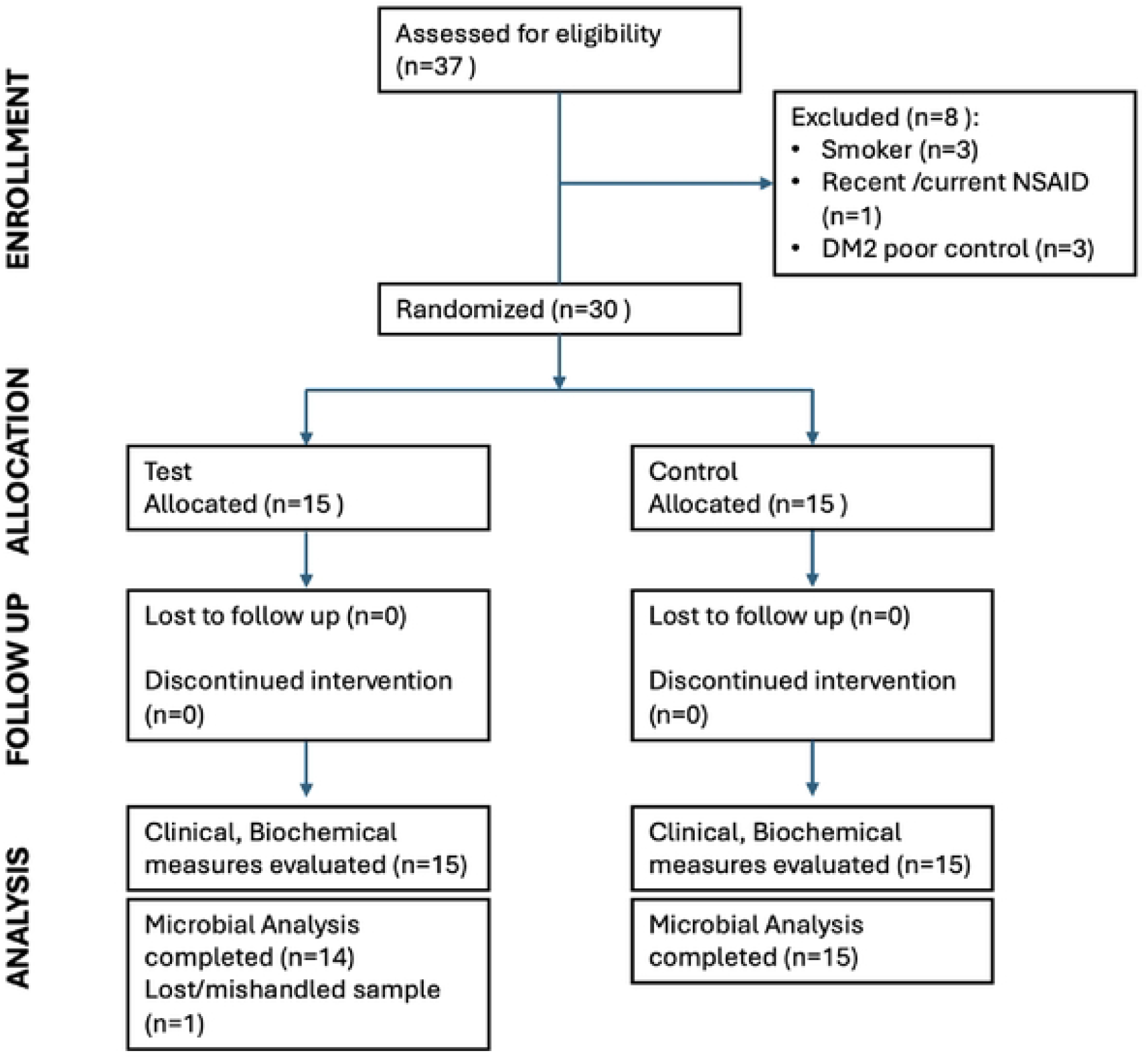
Clinical trial overview with recruitment, enrollment, allocation numbers, shown in CONSORT flow chart. Flow diagram illustrating participant screening, enrollment, allocation, follow-up, and analysis for the randomized, double-blind, placebo-controlled trial (ClinicalTrials.gov NCT06029283). A total of participants assessed for eligibility were screened according to inclusion and exclusion criteria. Eligible individuals were randomized in a 1:1 ratio to receive prebiotic nitrate chewing gum (n=15) or placebo chewing gum (n=15) for 21 days. All randomized participants completed the intervention and follow-up assessments. One baseline saliva sample in the test arm was lost during transport and excluded from microbiome analysis; all other participants were included in intention-to-treat analyses for clinical and biochemical endpoints. No participants discontinued due to adverse events.

**Table 1.**
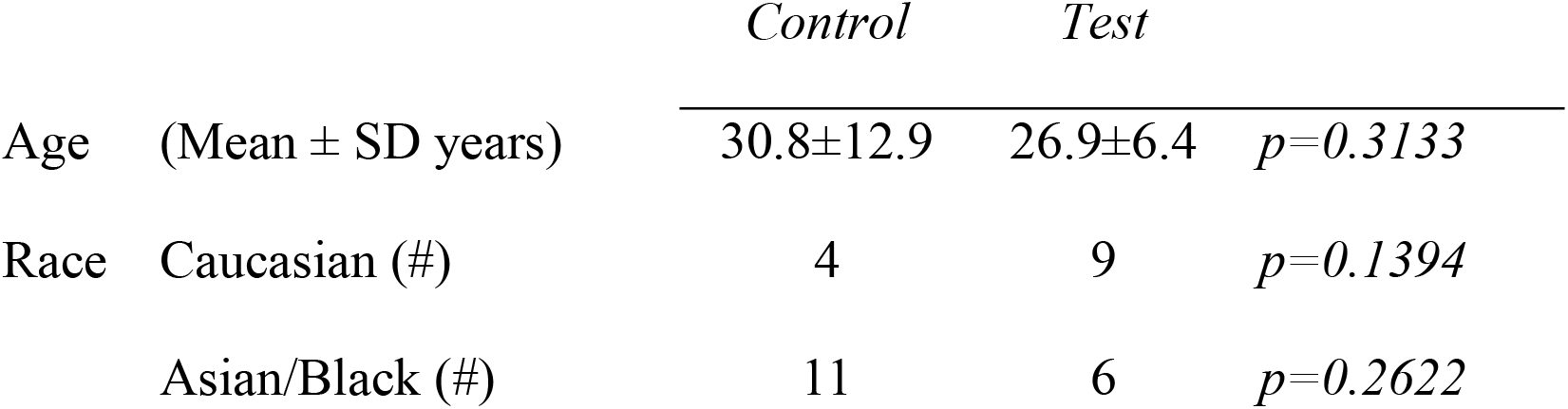
Baseline demographics of enrolled subjects. Subject compliance, measured as mean percentage of doses completed, showed no statistical difference between test (94.4%) and control (95.5%) participants. There were no reported adverse events, and all participants completed the study protocol visits.

### Effects of prebiotic nitrate functional gum on clinical measures and biochemical parameters

Following three weeks of intervention, the test group receiving prebiotic nitrate treatment demonstrated statistically significant changes in some indicators of oral biochemical and gingival health measures compared with the control group (**Table 2**)

**Table 2.**
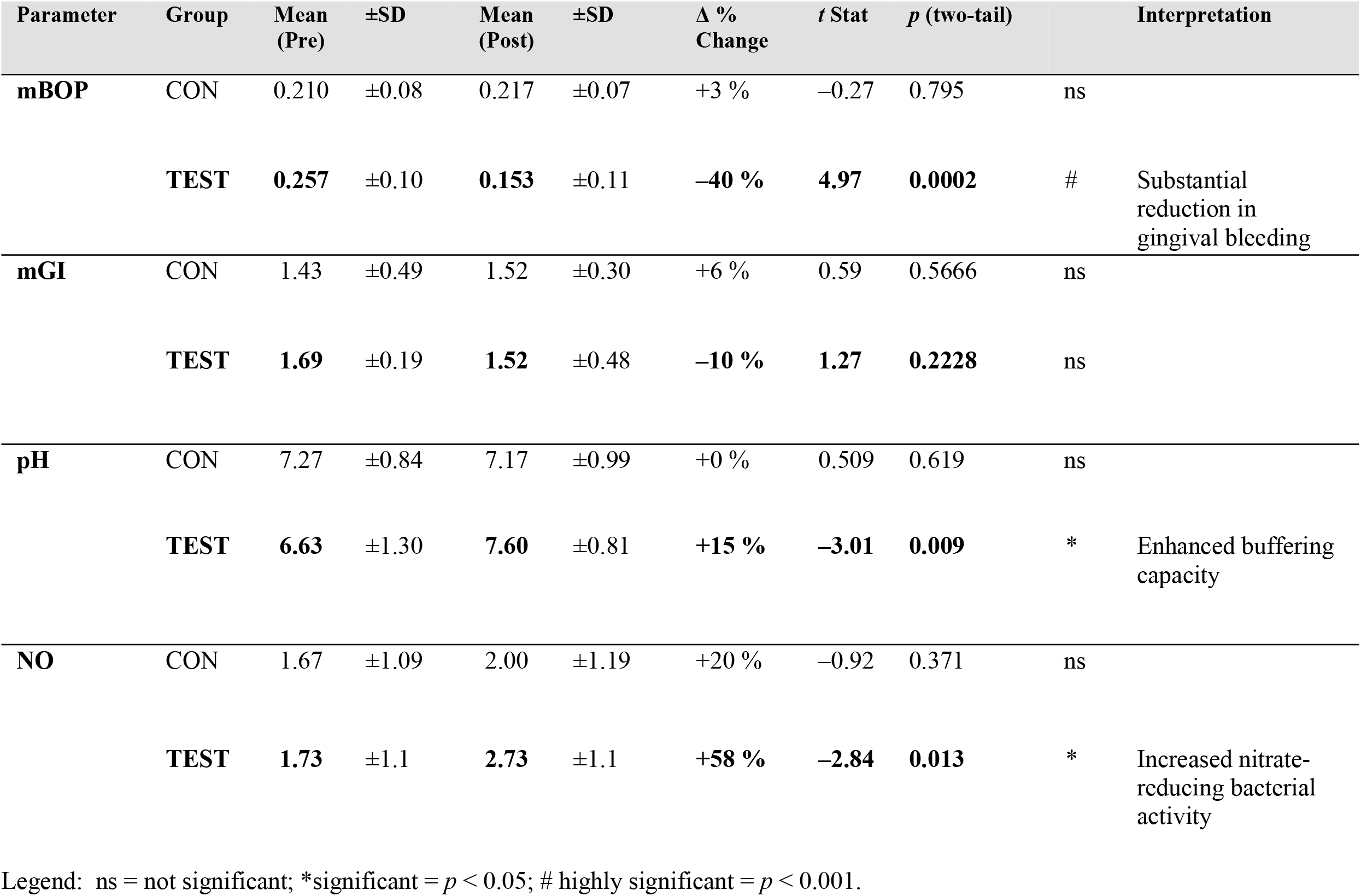
Pre- and post-treatment effects on mBOP, Salivary pH, Salivary NO, and mean Gingival Index. Mean Bleeding on Probing (mBOP) before and after 21-day intervention with prebiotic nitrate-chewing gum vs control gum. Mean mBOP (%) significantly decreased in the test group from 25.7% to 15.3% (p = 0.0002), indicating a reduction in clinical gingival inflammation. Control group showed no significant change (21.0% to 21.7%; p = 0.80). In test arm, salivary pH significantly increased from 6.63 ± 1.25 to 7.60 ± 0.79 (p = 0.009), indicating a shift toward a more alkaline oral environment. Salivary NOx, measured via semi-quantitative test strips (scale 1.0–5.0), increased in test arm from 1.73 ± 0.88 to 2.73 ± 1.10 (p = 0.0131), reflecting enhanced nitrate-reducing activity and nitric oxide bioavailability. Test strip scores were interpreted as follows: 1 = deficiency, 2 = low, 3 = target, 4 = optimal, 5 = elevated. Modified Gingival Index (mGI) decreased from 1.69 ± 0.20 to 1.52 ± 0.48; although this change was not statistically significant (p = 0.223), it indicates a trend toward reduced gingival inflammation.

Primary outcome mBOP score (**Figure 2**) decreased significantly from 0.26 ± 0.03 to 0.15 ± 0.02 (*–40%, p = 0.0002*), consistent with a visible reduction in erythema, edema, and tissue friability. In contrast, the control group mBOP showed no measurable benefit (0.21 → 0.22, p = 0.79).

**Figure 2.**
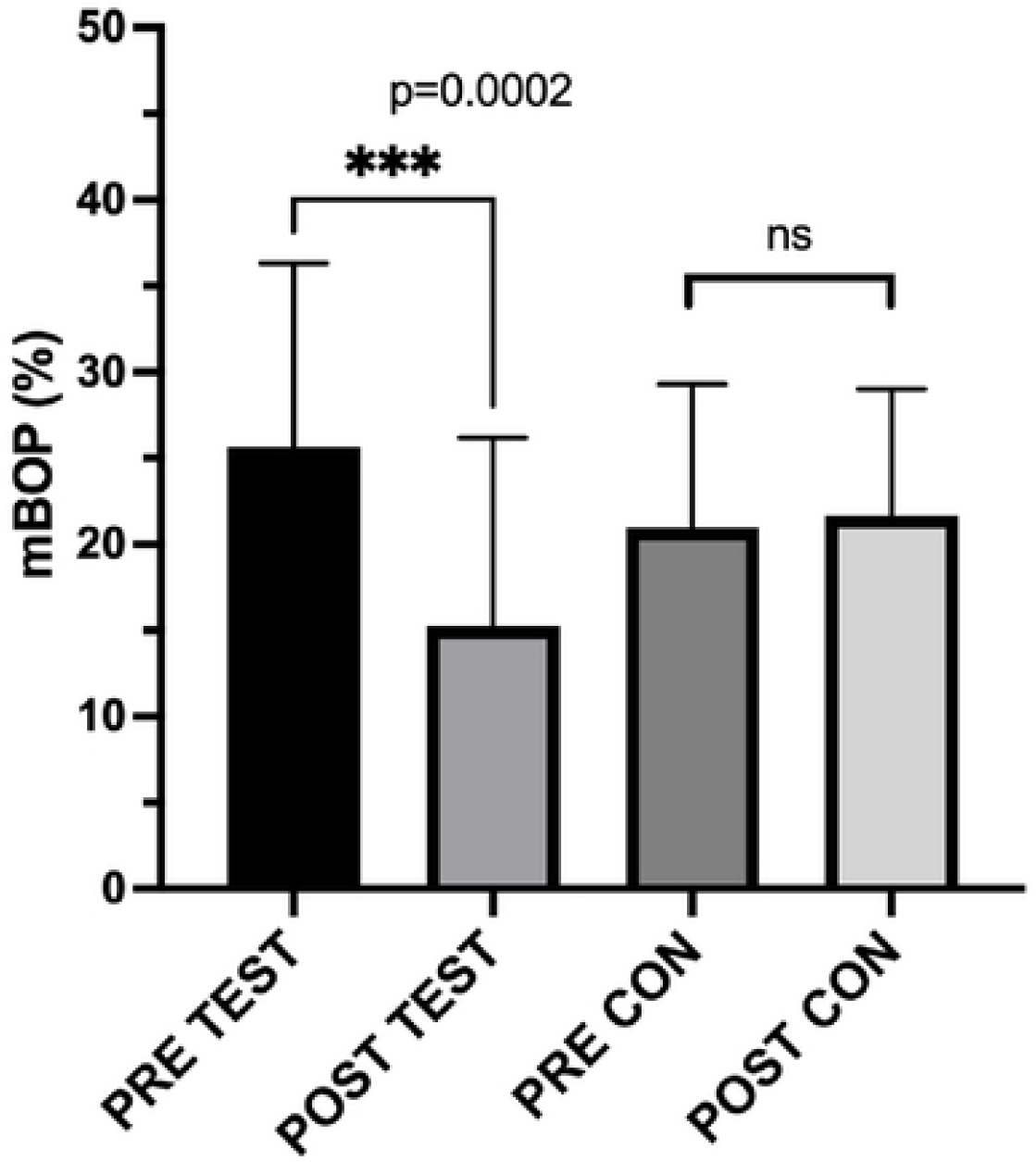
Pre- and post-treatmenteffects of test and control intervention on primary outcome metric, mean bleeding on probing (mBOP) Mean Bleeding on Probing (mBOP) before and after 21-day intervention with prebiotic nitrate-chewing gum vs control gum. Mean mBOP (%) significantly decreased in the test group from 25.7% to 15.3% (p = 0.0002), indicating a reduction in clinical gingival inflammation. Control group showed no significant change (21.0% to 21.7%; p = 0.80). Thin bars represent ± standard deviation (SD).

Changes in additional biochemical and clinical measures are presented in **Figure 3**. The modified Gingival Index (mGI) decreased non-significantly from 1.69 ± 0.14 to 1.52 ± 0.12 *p = 0.2228*) in the test group. A modest non-significant increase was observed associated with the control group mGI (1.43 → 1.52, p = 0.34).

**Figure 3.**
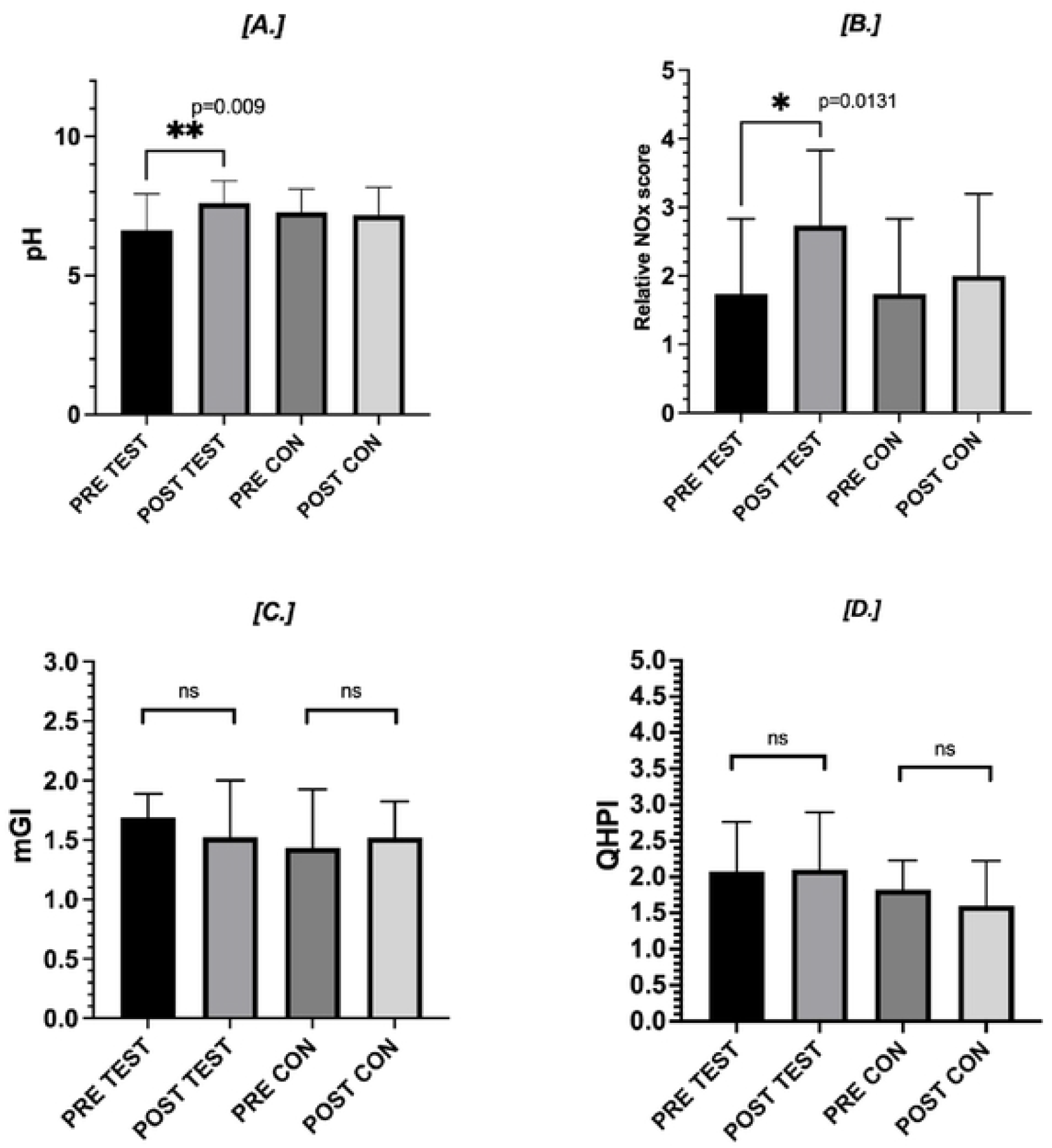
Pre- and post-treatmenteffects on Salivary pH, Salivary NO, mean Gingival Index and QHPI. Pre- and post-treatment changes in [A.] salivary pH, [B.] salivary nitric oxide, (NOx; nitrite), [C.] modified Gingival Index (mGI), [D.] QHPI, after 21 days of nitrate gum use (N = 15). In test arm, salivary pH significantly increased from 6.63 ± 1.25 to 7.60 ± 0.79 (p = 0.009), indicating a shift toward a more alkaline oral environment. Salivary NOx, measured via semi-quantitative test strips (scale 1.0–5.0), increased in test arm from 1.73 ± 0.88 to 2.73 ± 1.10 (p = 0.0131), reflecting enhanced nitrate-reducing activity and nitric oxide bioavailability. Test strip scores were interpreted as follows: 1 = deficiency, 2 = low, 3 = target, 4 = optimal, 5 = elevated. Modified Gingival Index (mGI) decreased from 1.69 ± 0.20 to 1.52 ± 0.48; although this change was not statistically significant (p = 0.223), it indicates a trend toward reduced gingival inflammation.

Test arm QHPI remained stable (2.08 to 2.10; p = 0.85), suggesting that improvements in gingival health occurred independently of visible changes in plaque biofilm levels. The QHPI score of the control arm also exhibited non-significant changes (2.02 to 1.93, p=0.2102).

Salivary pH increased from 6.63 ± 0.17 to 7.60 ± 0.12 (*p = 0.009*), representing a +15% rise toward a neutral environment that favors remineralization and suppresses acidogenic bacterial activity. In contrast, pH in the control group remained unchanged (7.27 → 7.17, *p = 0.62*).

Salivary nitric oxide (NO) levels rose markedly in the test group (1.73 → 2.73 µM, *+58%, p = 0.013*), reflecting enhanced bacterial nitrate-to-nitrite conversion and elevated NO bioavailability. The control group exhibited a minor, non-significant increase (1.67 → 2.00 µM, *p = 0.37*). This observation suggests that nitrate supplementation activated the oral nitrate–nitrite–NO pathway rather than spontaneous variation in salivary NO levels.

Collectively, these results suggest that local prebiotic nitrate formulated supplementation enhances oral nitric oxide metabolism and improves gingival health during 3-weeks. A clinically meaningful reduction in gingival inflammation, for a short-term period, is achieved by use of prebiotic nitrate formulated chewing gum.

### Analysis of microbiome composition before and after 21-day intervention with prebiotic nitrate functional gum

**Figure 4** present the species-level changes in mean relative abundance (Δ Post–Pre) for selected nitrate-reducing and pathogenic bacterial taxa following a 21-day intervention with dietary nitrate gum across 14 participants. Nitrate-reducing commensals—including *Rothia mucilaginosa, Neisseria subflava, Neisseria flavescens, Haemophilus parainfluenzae*, and *Neisseria sicca*—demonstrated robust cohort-wide increases in abundance, with *Rothia* and *Neisseria subflava* exhibiting the largest mean Δ values (+0.51 and +0.44, respectively). These findings are consistent with enhanced microbial nitrate metabolism and nitric oxide bioavailability observed in the salivary data.

**Figure 4.**
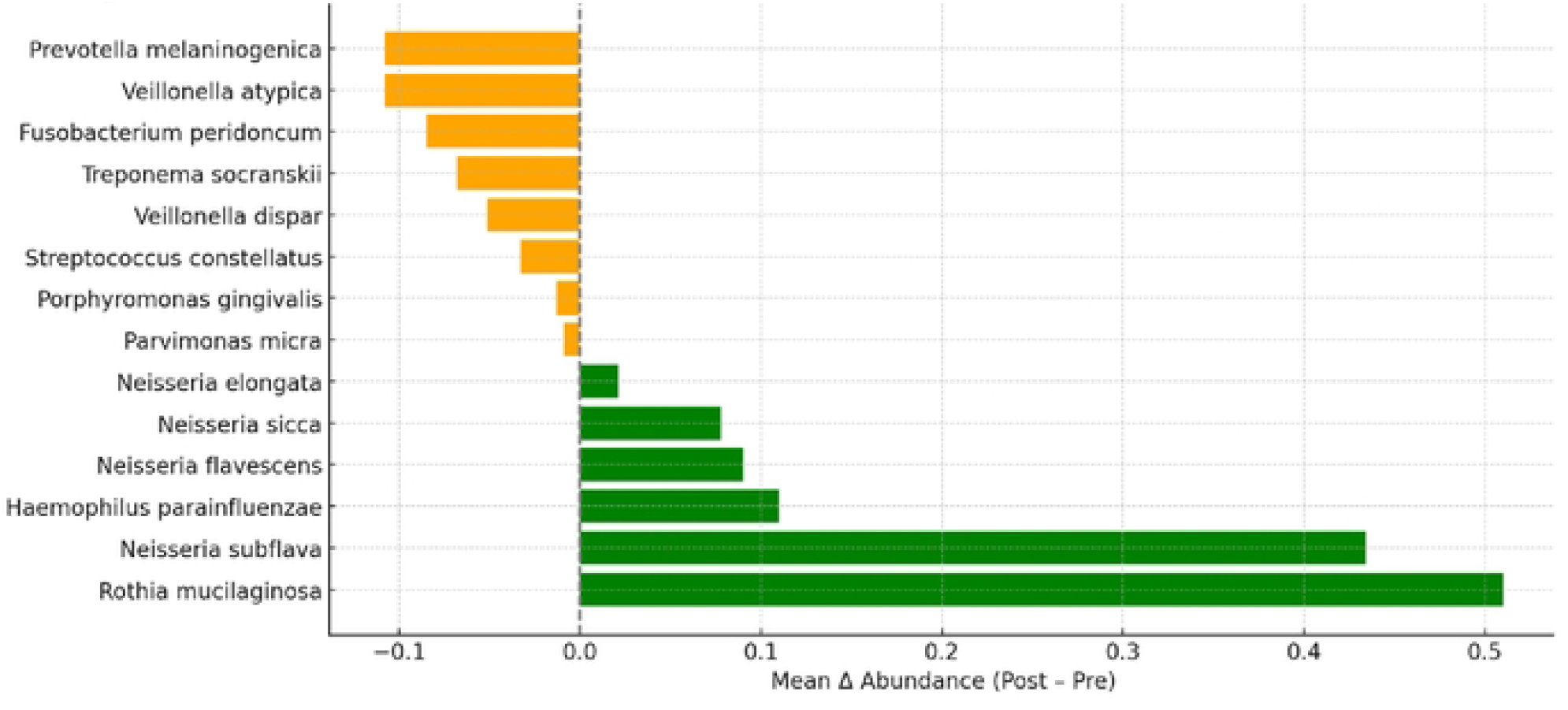
Microbiome shifts in nitrate-reducing and pathogenic-associated bacteria after 21-day prebiotic nitrate gum use. Bar graph showing mean change in relative abundance (Δ Post–Pre) for key nitrate-reducing (green bars) and pathogenic/inflammatory (orange bars) bacterial species across all 14 patients following 21 days of daily prebiotic nitrate gum use. Positive values indicate species that increased after treatment; negative values represent taxa that decreased. *Rothia mucilaginosa* and *Neisseria subflava* showed the greatest cohort-wide increases among nitrate reducers, while *Prevotella melaninogenica* and *Veillonella atypica* exhibited the largest decreases among NO-sensitive pathogenic taxa. Data reflect aggregated species-level changes from matched pre- and post-intervention microbiome sequencing.

Conversely, pathogenic and inflammation-associated taxa showed reproducible decreases across participants. *Prevotella melaninogenica, Veillonella atypica, Fusobacterium peridoncum, Treponema socranskii*, and *Porphyromonas gingivalis* each decreased in relative abundance, reflecting a shift away from dysbiosis. Notably, reductions were most pronounced in *Prevotella* and *Veillonella* species, genera previously associated with acidogenicity, inflammation, and nitric oxide sensitivity.

The directionality and magnitude of these changes support a hypothesis that localized slow-release nitrate supplementation can enrich NO-producing commensals while selectively suppressing periodontal and caries-associated pathogens. These compositional shifts parallel the improvements observed in gingival bleeding, salivary NO levels, and pH.

Bacterial species (**Table 3**) of interest were targeted by 16S rRNA identification based on prior evidence linking them to the nitrate–nitrite–NO pathway, or microbial dysbiosis. ^8-26^ Mean Δ values represent change in relative abundance units. Results reflect species-level assignments. The observed increases in *Rothia* and *Neisseria* species and decreases in *Veillonella, Prevotella*, and *Porphyromonas* support the hypothesis that nitrate gum promotes a eubiotic, NO-supportive oral environment.

**Table 3.** Mean Change in Abundance of Nitrate-Reducing and Pathogenic Microbial Species Following Prebiotic Nitrate Gum Intervention (Test Group) Species-level changes in oral microbiota following 21-day intervention with prebiotic nitrate functional gum. The table displays key nitrate-reducing commensals and inflammation-associated pathogenic species. For each, the mean change in relative abundance (Δ Post–Pre), standard deviation, patient prevalence, direction of change, and statistical significance are shown. P-values represent two-tailed paired t-tests across paired samples. Positive values reflect an increase in species abundance after treatment; negative values indicate a decrease. Directional arrows (↑/↓) denote consistent cohort-wide shifts. **\*Subjects** (n/14) indicates where n is # subjects with the identified species **and** majority shift in collected salivary sample.

| Species | Functional Category | Mean $\Delta$<br>Abundance | Standard<br>Deviation | Subjects*<br>(n/14) | Direction | p-<br>value |
| --- | --- | --- | --- | --- | --- | --- |
| *Rothia mucilaginosa* | Primary nitrate-reducer;<br>health-associated | 0.510 | 1.865 | 14/14 | ↑<br>Increased | 0.0014 |
| *Neisseria subflava* | Nitric oxide producer; anti-inflammatory | 0.434 | 1.564 | 14/14 | ↑<br>Increased | 0.0013 |
| *Haemophilus parainfluenzae* | Commensal nitrate-reducer;<br>systemic health linked | 0.110 | 0.424 | 14/14 | ↑<br>Increased | 0.0015 |
| *Neisseria flavescens* | Nitrate-reducer; associated<br>with vascular benefits | 0.090 | 0.202 | 14/14 | ↑<br>Increased | 0.0004 |
| *Neisseria sicca* | Oral commensal; nitrate<br>metabolism supportive | 0.078 | 0.283 | 14/14 | ↑<br>Increased | 0.0013 |
| *Neisseria elongata* | Robust nitrate-reducer; oral<br>nitric oxide pathway | 0.021 | 0.059 | 14/14 | ↑<br>Increased | 0.0007 |
| *Parvimonas micra* | Periodontitis-associated<br>pathogen | -0.009 | 0.061 | 12/14 | ↓<br>Decreased | 0.0380 |
| *Porphyromonas gingivalis* | Keystone periodontal<br>pathogen | -0.013 | 0.049 | 10/14 | ↓<br>Decreased | 0.0917 |
| *Streptococcus constellatus* | Inflammatory anaerobe;<br>dysbiosis indicator | -0.033 | 0.078 | 13/14 | ↓<br>Decreased | 0.2643 |
| Veillonella dispar | Acidogenic, NO-sensitive anaerobe | -0.051 | 0.202 | 13/14 | ↓<br>Decreased | 0.0836 |
| Treponema socranskii | Red-complex pathogen; inflammation-associated | -0.068 | 0.207 | 13/14 | ↓<br>Decreased | 0.1431 |
| Fusobacterium peridonticum | Inflammatory, periodontitis-linked | -0.085 | 0.138 | 11/14 | ↓<br>Decreased | 0.7197 |
| Veillonella atypica | NO-sensitive, acidogenic anaerobe | -0.108 | 0.274 | 14/14 | ↓<br>Decreased | 0.2207 |
| Prevotella melaninogenica | Pro-inflammatory; NO-sensitive genera | -0.108 | 0.303 | 13/14 | ↓<br>Decreased | 0.1727 |

### Stratified Microbiome Changes by mBOP Response

Stratified analysis of microbiome response by clinical improvement revealed key distinctions between high and low responders. Participants of the test group were grouped by gingival bleeding reduction (mBOP), with “high responders” defined as achieving ≥30% relative reduction (n = 7) and “low responders” as <30% improvement (n = 7). Microbial abundance outcomes were compared between these groups to identify response-associated patterns.

As shown in **Figure 5**, high responders demonstrated markedly greater enrichment of core nitrate-reducing taxa. *Rothia mucilaginosa* increased by +0.62 relative abundance units in high responders versus +0.23 in low responders (p = 0.017). Similarly, Neisseria elongata showed a +0.45 gain in high responders compared to +0.12 in low responders (p = 0.022). Other NO–promoting species, including *Neisseria subflava, Neisseria flavescens*, and *Haemophilus parainfluenzae*, were consistently elevated among high responders, suggesting stronger engagement of the nitrate–nitrite–NO pathway in those with greater clinical improvement.

**Figure 5.**
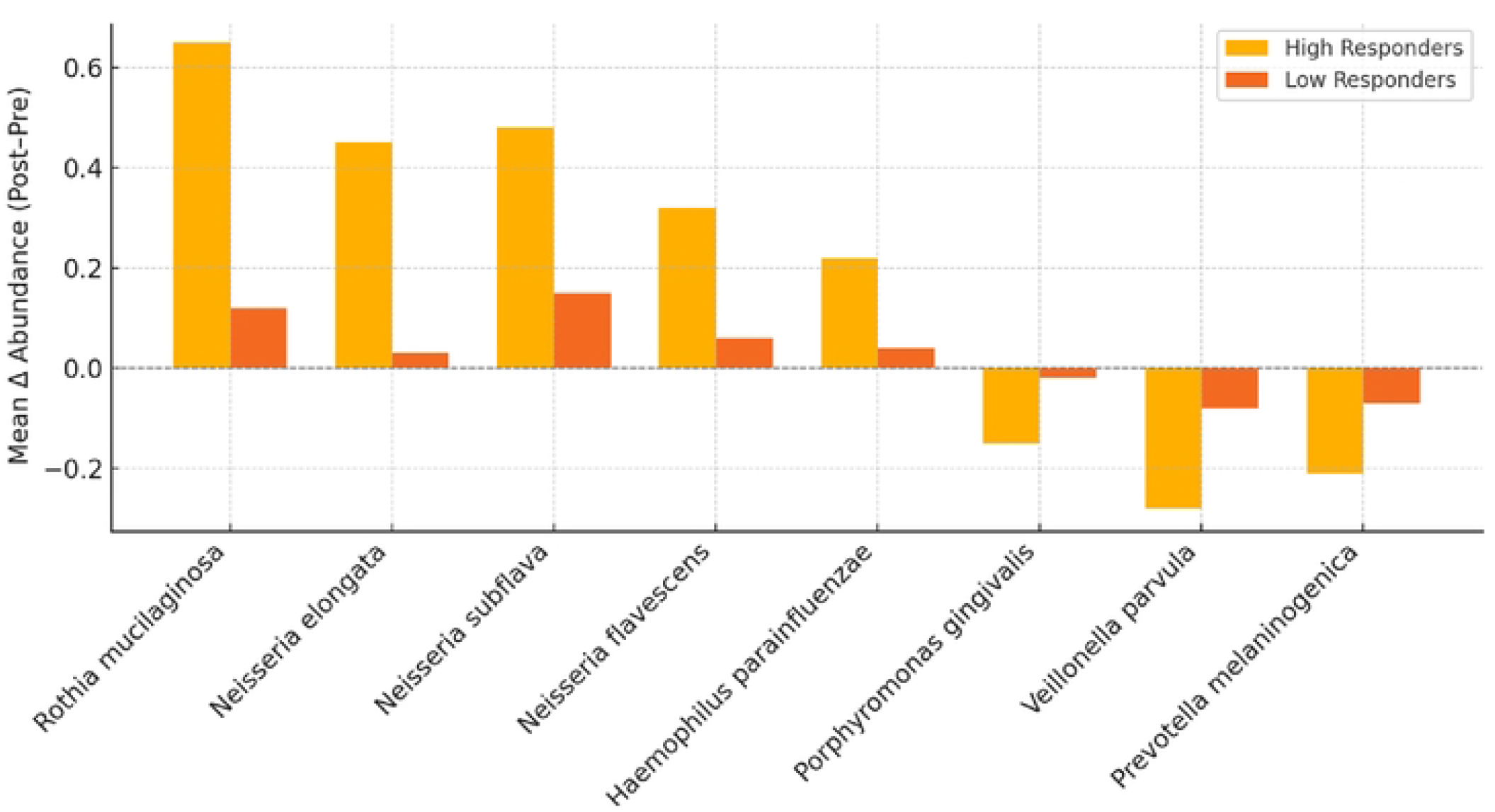
Microbial species-level abundance changes in high vs low clinical responders by mBOP to prebiotic nitrate gum intervention. Microbial shifts in high vs. low clinical responders stratified by mBOP reduction following nitrate gum intervention. Participants were classified as “high responders” (≥30% reduction in modified bleeding on probing [mBOP], n = 7) and “low responders” (<30% reduction, n = 7). The bar graph displays the mean change in relative abundance (Δ abundance: Post Pre) for five key nitrate-reducing species (e.g., *Rothia mucilaginosa, Neisseria spp*., *Haemophilus parainfluenzae*) and three pathogen-associated taxa (e.g., *Porphyromonas gingivalis, Veillonella parvula, Prevotella melaninogenica*). High responders exhibited consistently greater enrichment of beneficial nitrate-reducing bacteria and more pronounced suppression of pathogenic genera.

Conversely, high responders exhibited more pronounced suppression of pathogenic taxa. *Veillonella parvula* decreased by –0.25 in high responders versus –0.10 in low responders (p = 0.035), while *Porphyromonas gingivalis* declined by –0.18 compared to –0.03 (p = 0.048), reflecting enhanced NO-mediated pathogen clearance. *Prevotella melaninogenica*, another periodontitis-associated species, also declined more substantially in high responders, though not all changes reached statistical significance.

## Materials and Methods

### Study Design

The study was a randomized, double-blind, placebo-controlled trial (ClinicalTrials.gov ID: NCT06029283) conducted over a 21-day intervention period. The study protocol was prepared in accordance with the declaration of Helsinki and met GCP criteria; it was approved by the Institutional Review Board of the University of Maryland Baltimore (HP-00106795). All subjects enrolled had signed a written informed consent prior to inclusion. Allocation concealment was managed and maintained by study coordinator (HYK) and kept from both clinical evaluator (BY) and study team responsible for analysis (HJS, SG).

### Functional chewing gum

The functional chewing gum was formulated as a compressed, plant-based tablet designed for rapid buccal release during mastication by MyFitStrip LLC. Each tablet for the test arm contained 50 mg of food-grade prebiotic nitrate, derived from potassium.^21^ The gum was produced under GMP conditions using a low-heat direct compression method to preserve nitrate and polyphenol stability by Per Os Bioscience for MyFitStrip.^31,32^ Gum tablets for control (placebo) and test arms were stored and distributed in visually identical re-sealable pouches. Gum composition is detailed in the supplemental Appendix.

### Participants

Subjects were recruited and allocated using a computer-generated randomization into two groups (1:1 ratio). A test group received nitrate-containing chewing gum (∼0.989 mmol nitrate per piece). The control group received visually identical gum without prebiotic nitrate which showed no significant difference (data not shown). Baseline demographic parameters of age, sex, and race/ethnicity were collected.

#### Eligibility criteria

clinical presentation of gingival inflammation, as defined by modified gingival index (mGI) of 2.0 minimum; at least 20 natural teeth present; and no periodontitis, as determined by absence of radiographic interproximal bone loss.

#### Exclusion criteria

recent or current systemic antibiotic use, non-steroidal anti-inflammatory medication use in the past 14 days, use of anticoagulant therapy, recent periodontal therapy, current smoking, poorly controlled diabetes (HBA1c≥7.0%), and impaired ability to chew gum. In addition, subjects were not permitted to use oral rinses such as, chlorhexidine, essential oils, and cetylpyridinium chloride during the study.

All clinical assessment was completed at the University of Maryland School of Dentistry, Division of Periodontics clinic. All examination was completed by a single experienced dental practitioner (BY) not involved in allocation, nor directly handling of test/placebo gum packets. Intra-examiner calibration for collected clinical parameters was performed prior to the investigation (Kappa=0.91).

Enrolled subjects underwent clinical assessment, salivary microbial sampling and salivary nitric oxide and pH measurements at baseline (Day 0) and post-intervention (Day 21). Study subjects. were provided unlabeled pouches containing the needed units of gum for chewing test or placebo gum three times a day, for three weeks. Instructions were provided to utilize the gum for a minimum of 15 minutes at each dosage. Subjects agreed to refrain from use of any oral rinses containing chlorhexidine, essential oils or cetylpyridinium chloride. Subjects were otherwise asked to not change any existing oral hygiene behaviors or routines for the duration of the study. The subjects randomly received two surveys via email or SMS/text to monitor compliance durng the duration of the intervention. Each survey instance asked participant to acknowledge which of the scheduled three doses in the past 24 hours had been completed.

### Clinical Assessments

The primary outcome was mean Bleeding on Probing (mBOP), defined as the percentage of sites exhibiting bleeding upon gentle probing within 30 seconds. Each participant was assessed at six sites per tooth using a calibrated periodontal probe (UNC-15). mBOP was calculated as: mBOP (%) = Number of bleeding sites/Total number of sites probed ×100. Alternatively, mBOP may be expressed as a decimal equivalent. In addition, Quigley-hein plaque index (QHPI) and modified gingival index (mGI) were recorded at each time point.^33,34^

### Biochemical assessments

Salivary nitric oxide (NO) levels and oral pH were assessed using commercially available test strips (MyFitStrip LLC) designed for non-invasive, semi-quantitative colorimetric evaluation. These strips provide surrogate measurements of salivary nitrite as a proxy for bioavailable nitric oxide and allow concurrent assessment of oral cavity acidity via pH-sensitive indicators.

Participants were instructed to refrain from eating, drinking, or brushing their teeth for at least 30 minutes prior to sample collection to minimize variability due to recent oral activity. Each participant placed the absorption pad of the strip on the dorsal surface of the tongue for 5 seconds, then removed it and folded it over to compress against the nitrite-reactive pad containing Griess reagent.

For nitric oxide evaluation, the strip’s indicator region contains reagents that react with nitrite, an intermediate in the enterosalivary nitrate-nitrite-NO pathway, to produce a color change typically ranging from light pink (low NO availability) to deep magenta (high NO availability). Color intensity was compared to a manufacturer-provided reference scale and scored from 1 (depleted; <1 mg/L) to 5 (high; >250 mg/L).

pH assessment was simultaneously conducted using an adjacent pH-sensitive segment embedded on the same strip (MyFitStrip LLC). This zone changes color according to salivary acidity, corresponding to a pH scale typically ranging from 5.0 to 7.5. Readings were recorded by visual comparison to a calibrated color chart.

All measurements were performed under ambient conditions, and assessments were independently scored by trained personnel (BY) within 30 seconds of sample application to ensure optimal reactivity and reproducibility.^21,35,36^

### SALIVA Microbiome Profiling

Saliva samples were collected from participants at baseline (Day 0) and post-intervention (Day 21) using a commercially available saliva collection and microbiome profiling platform (Bristle Oral Health, San Francisco, CA). Participants were instructed to avoid eating, drinking, brushing, or using mouthwash for at least 30 minutes prior to sample collection. Each participant expectorated ∼1 mL of saliva into a sterile collection tube pre-filled with a DNA stabilization buffer (10mM Tris-HCL, 1 mM EDTA). Samples were processed by a Clinical Laboratory Improvement Amendments (CLIA) certified laboratory.

Microbial DNA was extracted using automated bead-beating and column purification methods. 16S rRNA gene sequencing targeting the V4 hypervariable region was performed using Illumina MiSeq (2×150 bp paired end reads). Raw sequencing data were demultiplexed, quality-filtered, and denoised using the DADA2 pipeline to generate amplicon sequence variants (ASVs). Taxonomic classification was assigned using a custom-trained naïve Bayes classifier against the Greengenes (13_8) or SILVA database (release 138), optimized for oral taxa. Downstream bioinformatic analyses, including relative abundance calculations, alpha diversity (Shannon index), and differential abundance testing, were conducted using QIIME 2.(19,20) In a secondary analysis, bacterial species were grouped into clinically relevant categories (e.g., nitrate reducers, periodontal pathogens, acidogenic taxa) for functional interpretation.^37,38^

### Statistical Analysis

Continuous and categorical baseline variables were compared using Student’s t-test and Fisher’s exact test, respectively. Paired t-tests evaluated within-group changes in mBOP, and independent t-tests assessed between-group differences. A two-tailed p < 0.05 was considered statistically significant. Data were analyzed using Python (v3.11, pandas/statsmodels) or GraphPad Prism (version 10.0).

## Discussion

This randomized, placebo-controlled trial demonstrates that a low-dose, prebiotic nitrate–enriched chewing gum formulation, delivered locally in a slow-release format, can reproducibly remodel the oral ecosystem toward a health-associated state within 21 days. Consistent and concordant improvements across clinical inflammation, salivary biochemistry, and microbiome composition support the concept that targeted nutritional modulation of oral biofilm function —rather than antimicrobial suppression— can meaningfully attenuate gingival inflammation.

The magnitude of clinical response was notable. Participants receiving prebiotic nitrate formulated gum experienced an approximately 40% reduction in mean bleeding on probing (mBOP), a sensitive and site-level marker of active gingival inflammation. This effect size is comparable to, or exceeds, outcomes reported in prior dietary nitrate interventions using beetroot juice or nitrate-rich foods, despite the substantially lower total nitrate dose used here. The absence of a statistically significant change in modified Gingival Index (mGI), despite a consistent directional trend, likely reflects methodological differences between indices. Whereas mGI relies on partial-mouth visual scoring, mBOP captures full-mouth inflammatory responses and may therefore be more responsive to early or subtle shifts in gingival immune tone.

At the microbial level, nitrate gum administration produced a strikingly consistent enrichment of nitrate-reducing, health-associated taxa—including *Rothia mucilaginosa* and multiple *Neisseria* species—across nearly all participants in the intervention arm. These organisms are central to the oral nitrate–nitrite–nitric oxide (NO) pathway and contribute to local vasodilation, immune modulation, pH buffering, and ecological stability.^9,11-26^ Importantly, the magnitude and consistency of these shifts parallel those reported in higher-dose systemic nitrate studies, indicating that sustained local nitrate availability—rather than total nitrate load—is sufficient to restructure oral biofilm function.

Concomitant reductions were observed in nitrate-sensitive, inflammation-associated taxa, including *Prevotella melaninogenica, Fusobacterium nucleatum, Veillonella* spp., *Porphyromonas gingivalis, Treponema socranskii*, and *Streptococcus constellatus*. These organisms are well recognized contributors to proteolytic metabolism, tissue breakdown, and inflammatory signaling within dysbiotic periodontal biofilms. The observed suppression of these taxa is consistent with prior in vitro and ex vivo studies demonstrating that nitrate-derived nitrite and NO exert selective inhibitory pressure on anaerobic and proteolytic species, while sparing or promoting facultative nitrate reducers.^9,13^ In parallel, increases in salivary pH may further disfavor acidogenic and proteolytic taxa, reinforcing ecological rebalancing.

Stratified responder analyses provide additional mechanistic insight. Participants achieving ≥30% reductions in mBOP exhibited markedly greater enrichment of nitrate-reducing commensals—particularly *Rothia mucilaginosa, Neisseria elongata, Neisseria subflava, Neisseria flavescens*, and *Haemophilus parainfluenzae*—alongside sharper declines in established periodontal pathogens. These findings support a microbiome-mediated mechanism of action in which clinical benefit is directly linked to the magnitude of nitrate-reducing capacity gained within the biofilm. Beyond serving as functional effectors, these taxa may represent predictive biomarkers of nitrate responsiveness, suggesting opportunities for microbiome-informed personalization of therapy.

Our findings align closely with prior clinical and experimental work. Jockel-Schneider et al. reported improved gingival inflammation and enrichment of *Rothia* and *Neisseria* following nitrate-rich juice supplementation, while Mazurel et al. demonstrated dose-dependent reductions in dysbiosis and enhanced nitrate-reduction activity in ex vivo subgingival biofilms exposed to physiologically relevant nitrate concentrations.^17,18^ Notably, Mazurel et al. observed substantial ecological improvements even at 5 mM nitrate, reinforcing the translational relevance of the low-dose, localized approach employed here.

Taken together, these data support two complementary and reinforcing mechanisms by which localized prebiotic nitrate attenuates gingival inflammation. First, nitrate-derived NO likely modulates the local redox environment, reducing oxidative stress and dampening excessive leukocyte activation and cytokine release, thereby promoting resolution of inflammation. Second, nitrate acts as a selective ecological driver that enriches nitrate-reducing commensals and suppresses dysbiosis-associated anaerobes, restoring a functionally balanced biofilm. Prior work by Li et al., Mitsui et al., and Sanchez et al. similarly demonstrated nitrate-associated reductions in acidogenic and anaerobic taxa alongside enhanced microbial stability. ^5,28,29,30^ These effects are further supported by the emerging “oral nitrate hang-time” hypothesis, which posits that prolonged oral exposure amplifies both microbial and vascular benefits.^21^

While this study was designed as a short-term proof-of-concept intervention, the consistency of responses across participants and endpoints strengthens confidence in the biological robustness of the observed effects. The modest cohort size and 21-day duration were sufficient to detect coordinated shifts in inflammation, NO bioavailability, and microbial ecology, underscoring the sensitivity of the oral ecosystem to targeted nitrate modulation. Longer-term studies will be valuable to define durability of response, optimal dosing strategies, and potential synergistic combinations with probiotic nitrate-reducing strains. Importantly, the present findings establish a strong mechanistic and translational foundation upon which such studies can be built.

In conclusion, this study demonstrates that low-dose, slow-release nitrate chewing gum or lozenge can reproducibly enhance oral NO bioavailability, remodel the oral microbiome toward eubiosis, suppress dysbiosis-associated taxa, and improve gingival inflammation. The non-invasive, user-friendly format and low-cost production potential of this prebiotic approach position it as a scalable strategy for preventive oral care. More broadly, these findings support a paradigm shift toward biofilm-directed nutritional interventions that restore microbial function rather than indiscriminately suppress microbial communities.

## Data Availability

Data collected for the study will be made available and shared with others though contact at the following address. The data will be shared with researchers who want to do additional analysis of the data and therefore the data will be shared after approval of a proposal, and with a signed data access agreement.

## Trial registration

ClinicalTrials.gov Identifier: NCT06029283, Study Registration Submitted 2023-08-31, Study Completion 2025-08-01. The *CONSORT* protocol was approved by the Institutional Review Board of the University of Maryland, Baltimore (HP-00106795). All participants provided written informed consent. The study complied with the Declaration of Helsinki and Good Clinical Practice.

## Author Contributions

Conceptualization, H.J.S. and S.J.G.; experiment and investigation, H.J.S. and S.J.G.; Resources, Methodology, data curation, H.J.S. and S.J.G., B.Y, and H.Y.K, writing-original draft preparation, H.J.S. and S.J.G.; writing-review and editing, H.J.S. and S.J.G.; funding acquisition, H.J.S. and S.J.G., R.E. All authors have read and approved the final version of the manuscript. H.J.S. and S.J.G., B.Y, and H.Y.K. have accessed and verified the underlying data.

## Reference

1. Lang NP, Joss A, Orsanic T, Gusberti FA, Siegrist BE. Bleeding on probing. A predictor for the progression of periodontal disease? J Clin Periodontol. 1986 July;13(6):590–6.

2. Chapple ILC, Mealey BL, Van Dyke TE, Bartold PM, Dommisch H, Eickholz P, et al. Periodontal health and gingival diseases and conditions on an intact and a reduced periodontium: Consensus report of workgroup 1 of the 2017 World Workshop on the Classification of Periodontal and Peri-Implant Diseases and Conditions. J Clin Periodontol. 2018 June;45 Suppl 20:S68–77.

3. Nazir M, Al-Ansari A, Al-Khalifa K, Alhareky M, Gaffar B, Almas K. Global Prevalence of Periodontal Disease and Lack of Its Surveillance. Scientific World Journal. 2020;2020:2146160.

4. Lamont RJ, Koo H, Hajishengallis G. The oral microbiota: dynamic communities and host interactions. Nat Rev Microbiol. 2018 Dec;16(12):745–59.

5. Li Y, He X, Luo G, Zhao J, Bai G, Xu D. Innovative strategies targeting oral microbial dysbiosis: unraveling mechanisms and advancing therapies for periodontitis. Front Cell Infect Microbiol. 2025;15:1556688.

6. Lundberg JO, Weitzberg E. Physiology of nitrate and nitrite in health and disease. Nat Rev Urol. 2014;11(12):703–714. doi:10.1038/nrurol.2014.325.

7. Green SJ. Nitric oxide in mucosal immunity. Nature Medicine 1, 515–517 (1995).

8. Siervo M, Lara J, Ogbonmwan I, Mathers JC. Inorganic Nitrate and Beetroot Juice Supplementation Reduces Blood Pressure in Adults: A Systematic Review and Meta-Analysis. J Nutr. 2013 June;143(6):818–26.

9. Rosier BT, Buetas E, Moya-Gonzalvez EM, Artacho A, Mira A. Nitrate as a potential prebiotic for the oral microbiome. Sci Rep. 2020 July 30;10(1):12895.

10. Alhulaefi SS, Watson AW, Ramsay SE, Jakubovics NS, Matu J, Griffiths A, et al. Effects of dietary nitrate supplementation on oral health and associated markers of systemic health: a systematic review. Crit Rev Food Sci Nutr. 2025;65(14):2813–28.

11. Rosier BT, Takahashi N, Zaura E, Krom BP, Martínez-Espinosa RM, van Breda SGJ, et al. The Importance of Nitrate Reduction for Oral Health. J Dent Res. 2022 July 1;101(8):887–97.

12. Nambu T, Wang D, Mashimo C, Maruyama H, Kashiwagi K, Yoshikawa K, et al. Nitric Oxide Donor Modulates a Multispecies Oral Bacterial Community—An In Vitro Study. Microorganisms. 2019 Sept;7(9):353.

13. Fujii, A. et al. Enhanced dominance of nitrate-reducing bacteria using a combination of nitrate and erythritol in in vitro cultured oral biofilm. J. Oral Microbiol. 17, 2526069 (2025).

14. Akatsu, T. et al. Characteristics of subgingival plaque microbiome in Japanese older adults with healthy gingiva. J. Clin. Periodontol. 52, e14192 (2025).

15. Fukuda, S. et al. Commensal *Neisseria* inhibit *Porphyromonas gingivalis* invasion of gingival epithelial cells. Oral Health Prev. Dent. 22, 609–616 (2024).

16. Moran SP, Rosier BT, Henriquez FL, Burleigh MC. The effects of nitrate on the oral microbiome: a systematic review investigating prebiotic potential. J Oral Microbiol. 16(1):2322228.

17. Jockel-Schneider Y, Schlagenhauf U, Stölzel P, Goßner S, Carle R, Ehmke B, et al. Nitrate-rich diet alters the composition of the oral microbiota in periodontal recall patients. J Periodontol. 2021 Nov;92(11):1536–45.

18. Mazurel D, Carda-Diéguez M, Langenburg T, Žiemytė M, Johnston W, Martínez CP, et al. Nitrate and a nitrate-reducing Rothia aeria strain as potential prebiotic or synbiotic treatments for periodontitis. Npj Biofilms Microbiomes. 2023 June 17;9(1):40.

19. Bondonno CP, Liu AH, Croft KD, Ward NC, Puddey IB, Woodman RJ, et al. Short-Term Effects of a High Nitrate Diet on Nitrate Metabolism in Healthy Individuals. Nutrients. 2015 Mar;7(3):1906–15.

20. Hyde ER, Andrade F, Vaksman Z, Parthasarathy K, Jiang H, Parthasarathy DK, et al. Metagenomic Analysis of Nitrate-Reducing Bacteria in the Oral Cavity: Implications for Nitric Oxide Homeostasis. PLOS ONE. 2014 Mar 26;9(3):e88645.

21. Green J, Green SJ. Topical prebiotic nitrate: can extending the “hang-time” in the mouth improve oral-vascular health outcomes? NPJ Biofilms Microbiomes. 2024 July 18;10(1):57.

22. Goh, C. E. et al. Association between nitrate-reducing oral bacteria and cardiometabolic outcomes: results from ORIGINS. J. Am. Heart Assoc. 8, e013324 (2019).

23. Velmurugan S, Gan JM, Rathod KS, Khambata RS, Ghosh SM, Hartley A, et al. Dietary nitrate improves vascular function in patients with hypercholesterolemia: a randomized, double-blind, placebo-controlled study. Am J Clin Nutr. 2016 Jan;103(1):25–38.

24. Vanhatalo A, Blackwell JR, L’Heureux JE, Williams DW, Smith A, van der Giezen M, et al. Nitrate-responsive oral microbiome modulates nitric oxide homeostasis and blood pressure in humans. Free Radic Biol Med. 2018 Aug 20;124:21–30.

25. Chai X, Liu L, Chen F. Oral nitrate-reducing bacteria as potential probiotics for blood pressure homeostasis. Front Cardiovasc Med. 2024 Apr 4;11:1337281.

26. Rosier BT, Mazurel D, de Jager M, Zaura E, Loos BG, Crielaard W. Nitrate reduction capacity of the oral microbiota is impaired in periodontitis. npj Biofilms Microbiomes. 2023;9(1):64. doi:10.1038/s41522-023-00428-7.

27. Doel JJ, Hector MP, Amirtham CV, Al-Anzan LA, Benjamin N, Allaker RP. Protective effect of salivary nitrate and microbial nitrate reductase activity against caries. Eur J Oral Sci. 2004 Oct;112(5):424–8.

28. Silva Mendez LS, Allaker RP, Hardie JM, Benjamin N. Antimicrobial effect of acidified nitrite on cariogenic bacteria. Oral Microbiol Immunol. 1999 Dec;14(6):391–2.

29. Mitsui T, Fujihara M, Harasawa R. Salivary nitrate and nitrite may have antimicrobial effects on Desulfovibrio species. Biosci Biotechnol Biochem. 2013;77(12):2489–91.

30. Sánchez GA, Miozza VA, Delgado A, Busch L. Total salivary nitrates and nitrites in oral health and periodontal disease. Nitric Oxide Biol Chem. 2014 Jan 30;36:31–5.

31. Bobe K, Suryawanshi Y, Gomase V, Kachhi M, Bobade C. Formulation Consideration of Medicated Chewing Gum: A Review. Recent Adv Drug Deliv Formul. 2024;18(2):100–9.

32. Antony A, Farid M. Effect of Temperatures on Polyphenols during Extraction. Appl Sci. 2022 Jan;12(4):2107.

33. Lobene RR, Weatherford T, Ross NM, Lamm RA, Menaker L. A modified gingival index for use in clinical trials. Clin Prev Dent. 1986;8(1):3–6.

34. Löe H. The Gingival Index, the Plaque Index and the Retention Index Systems. J Periodontol. 1967;38(6):Suppl:610–616.

35. McDonagh STJ, Wylie LJ, Webster JMA, Vanhatalo A, Jones AM. Influence of dietary nitrate food forms on nitrate metabolism and blood pressure in healthy normotensive adults. Nitric Oxide. 2018;72:66–74. doi:10.1016/j.niox.2017.12.001.

36. Houston MC, Minich DM, Sinatra ST, Kahn JK, Green SJ Effects of a nitric oxide–supporting dietary supplement on blood pressure and salivary nitric oxide: an open-label clinical trial. Cureus. 2023;15(3):e36063. doi:10.7759/cureus.36063.

37. Regueira-Iglesias A, Balsa-Castro C, Blanco-Pintos T, Tomás I. Critical review of 16S rRNA gene sequencing workflow in microbiome studies: From primer selection to advanced data analysis. Mol Oral Microbiol. 2023;38(5):347–99.

38. Bolyen E, Rideout JR, Dillon MR, Bokulich NA, Abnet CC, Al-Ghalith GA, et al. Reproducible, interactive, scalable and extensible microbiome data science using QIIME 2. Nat Biotechnol. 2019 Aug;37(8):852–7.

